# System Dynamics Model of Possible Covid-19 Trajectories Under Various Non-Pharmaceutical Intervention Options in a Low Resource Setting

**DOI:** 10.1101/2020.10.06.20204487

**Authors:** Kivuti-Bitok W Lucy, Momodu S.Abiodun, Cheptum Jebet Joyce, Kimemia Fredrick, Gichuki Isaac, Ngune Irene

**Affiliations:** School of Nursing sciences, University of Nairobi, Kenya; Centre for Energy Research and Development, Obafemi Awolowo University, Nigeria; Health System Management AssociationKenya; School of Nursing, Midwifery and Paramedicine; Faculty of Health Sciences; Curtin University, Australia

## Abstract

We present a population-based System Dynamics Model (SDM) of possible Covid-19 trajectories under various intervention options in the uniqueness of Kenya. We developed a stock and flow based SDM. We parametrized the SDM using published data and where data was not available, expert opinion was sought. Following validation test, the model was simulated to determined possible outcomes of non-pharmaceutical interventions in management of Covid-19. We simulate the possible impact of; social distancing, quarantining, curfew and cross-county travel restriction, lockdown of selected cities in Kenya and quarantining. We varied interventions in terms of start dates, duration of implementation and effectiveness of the interventions. We estimated the outcomes in terms of number of possible infections, recoveries and deaths. With the current state of interventions, we estimated a peak of Covid-19 in September 2020 with an estimated 13.5 Million Covid-19 cases and 33.8 thousand deaths in Kenya. The largest possible reduction in infections and mortality was achievable through increase in the effectiveness of the interventions. The suggested interventions would delay the epidemic peak of Covid-19 to between late Nov 2020 and early December 2020 with an estimated13M cases a 500 thousand reduction in Covid-19 cases and 32.4 deaths (a reduction in 1400 deaths). We conclude that SDM enables understanding of the complexity and impact of different interventions scenarios of Covid-19 in Kenya.

## 1.0 BACKGROUND

Covid-19 a disease caused by severe acute respiratory syndrome coronavirus 2 (SARS-CoV-2)[1][2]continues to ravage the globe with a risk of catastrophic effect if no mitigation measures are put in place including an impending of the overwhelming of health care facilities worldwide. WHO declared Covid-19 an pandemic[3]. It is estimated that if no effective interventions are put in place, there will be approximated 7 billion infections and 40 million deaths worldwide [4]. Worldwide, consulted effort to have an in-depth understanding of the current and future trajectories of covid-19 within various interventions [5][6].Covid-19 is a great threat to the healthcare systems of the world and especially in Sub-Saharan Africa[1][1].

Kenya a sub-Saharan country with fragile health care system reported its first case of Covid-19 on the 13^th^ of March 2020 [7]. The case was an import in to the country. Kenya confirmed community transmission on the 30^th^ of March 2020 [8]. By 19^th^ August 2020, Kenya had reported - 30636 confirmed Covid-19 cases, 17,368 recoveries and 487 deaths (https://www.coronatracker.com/country/kenya/) Kenya has implemented a number of strategies in managing Covid-19. These include social distancing, curfew, quarantining, closing of social facilities, sanitizing and basic hygiene measures, reduction of public vehicles passengers by 60%, restriction of hospital visits and [9] modified lockdown (border closure) of five (Nairobi, Mombasa, Kilifi, Mandera and Kwale) main hotspot counties in the country [10], closure of schools, bars, and religious gathering among other measures.

Our objective was establishing the impact of these locally adapted non-pharmaceutical measures which are aimed at managing the spread of Covid-19 and flattening the curve in tandem with global trends [11]. With an expected exponential growth of Covid-19 in the country, it is important to gain an in-depth understanding of the interplay of different variables in the spread of the disease, their interactions and the probable impact of different intervention options at the population level.

One tool that can be applied in developing an in-depth understanding of the Covid-19-Transmission and impact of mitigation at the general population level is System Dynamics modelling (SDM)[12]. Modeling of outbreaks that threatens public health system has been found to be highly valuable to answering “high-stakes policy questions” [13], especially for developing countries like Kenya. System Dynamics modeling approach has been used to demonstrate both qualitative and quantitative varied options to managing such pandemic as COVID-19 [14]. SDM has been advocated as a tool predicting the number of new cases as well as identification of best measures to mitigate SARS-CoV-2 transmission [11].

### 1.1 SYSTEM DYNAMICS MODELLING (SDM)

Developed by Jay Forrester in the late 1950s [15], [16] SDM (the origin of current whole systems thinking) is a differential equations-based model that involves a number of steps. The activities are ‘(1) problem identification and definition, (2) system conceptualization, (3) model formulation, (4) model testing and evaluation, (5) model use, implementation and dissemination, and (6) design of learning strategy/infrastructure’[17].

Mental models of dynamic wicked problems such as Covid-19 are presented using Causal Loop Diagrams (CLD). These are further developed into a comprehensive Computerized model using software such as Stella^®^ and Vensim^®.^ The variables identified in these CLD are translated in terms of Stocks (depicting variables that accumulate in number) and flows between the stocks as well as the information that determines the value of the flows (converter variables)[18][19]. Feedback effects and delays are a key component of SDM. Differential equations are the main drivers of the model.

*In silico* experimentations, which combine findings from literature and computerized mathematical models, allow vast numbers of experiments that may produce more accurate results that gives room for hypothesis generation [20][21][22]. Computerized experimentations are cost effective and are less time-consuming alternative to expensive real time laboratory and clinical experimentation. Simulation using computer software enables study of systems behavior over time and supports *in silico* policy analysis. SDM relies on existing qualitative and quantitative data, and where data is not available, expert opinion is sought [23] The CLDs, stocks and flows provide a common language that can be easily understood by a wide range of stakeholders.

SDM has been recommended in analysis and understanding of the impact of different interventions in management of Covid-19 [24][12][25] [26]. The effect of quarantine periods on contacts and deaths in Covid-19 has been modelled [26]. At the National level, SDM has been recommended as a versatile tool in decision making in population-based models [12] in regards to quarantine, social distancing, delivery of testing, hospital capacity, staffing, resource mobilization well as health and wellbeing of the patient. At the global level SDM can be utilized in better understanding of the impact of global quarantine [12].SDM is generally used for strategic decisions affecting the whole population.

Studies applying SDM have been conducted in a variety of settings. Few if any SDM of Covid-19 studies have been done in Sub-Sharan Africa. We adopted Susceptible, Infected and Removed(SIR)structure [27] and hence splits the study population into mutually exclusive groups, subgroups and compartments. In line with other modelling studies on epidemic and pandemics[28][4]we separated the susceptible to include the exposed and the removed to include the recovered and the dead. Thus we adapted a Susceptible(persons who have not contracted but have potential to contract the virus), Exposed(Persons who have come into contact with an infected person and may or may not have contracted the virus and are at the same time asymptomatic), Infected(persons who have contracted the disease, they may or may not be symptomatic and may infect others), Recovered (persons who had been infected with the virus and whose infection has cleared and may no longer infect others)and Death(persons who succumb to the viral infection) (SEIRD) model to simulate Covid-19 trajectory under different scenarios in Kenya.

Appendix 1. Map of Kenya Showing the Various Covid19 distribution per county and the hotspot counties.

## 2.0 METHODS

### 2.1 Model Structure and Extension

Using the general structure of SEIRD model, we outlined the progression of Covid-19 through different pathways under different non pharmaceutical interventions. Using Stella Architect Version^R^ 2.0 we outlined the stocks, flows, converters and connectors in accordance to the practice of System Dynamics [29][16][19]. The basic structures used to build the model are illustrated in Fig 1.

**Figure 1:**
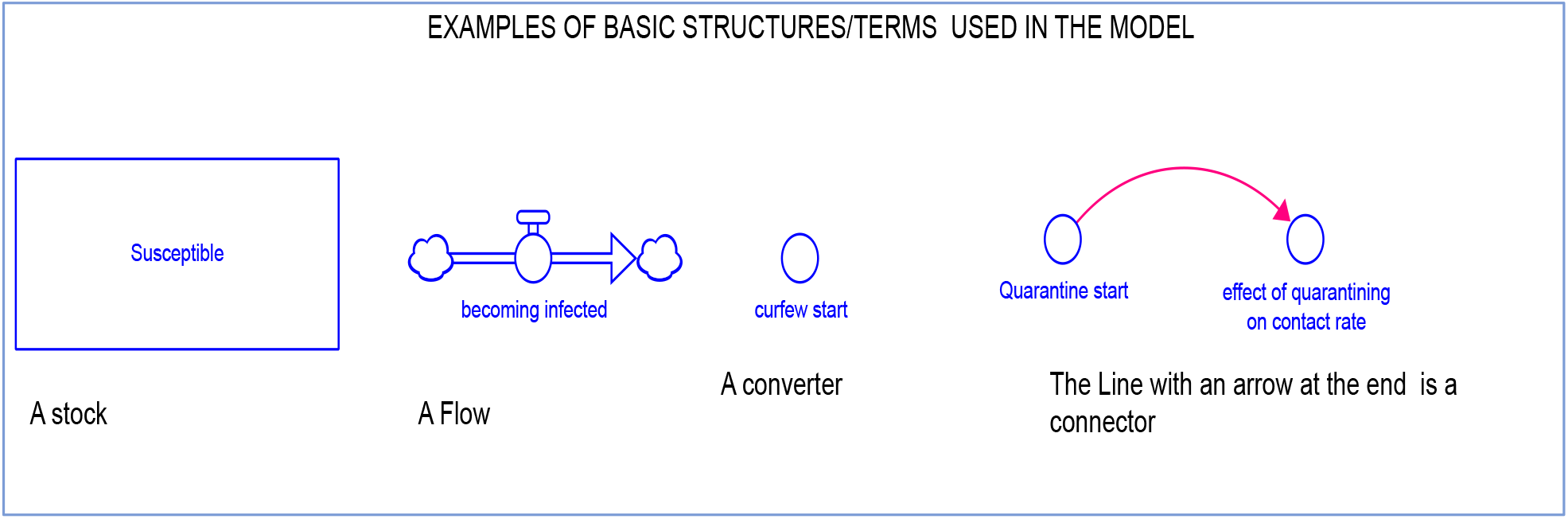
Basic structures and terms used in SDM

The general structure of the model presented as stocks and flows diagram is shown in Figure 2. Stocks represent variables that accumulate and are measured by levels. In this study, the stocks represent the number of people in each state regarding Covid-19 thus one may be susceptible, exposed, infected, recovered or dead. The flows represent the movement from one state to the next at a given time. Transit from one state (stock) to another is guided by a general set of equations. The mathematical model representing the studied problem can be obtained from the equation model view of supplementary file 1.

**Fig 2:**
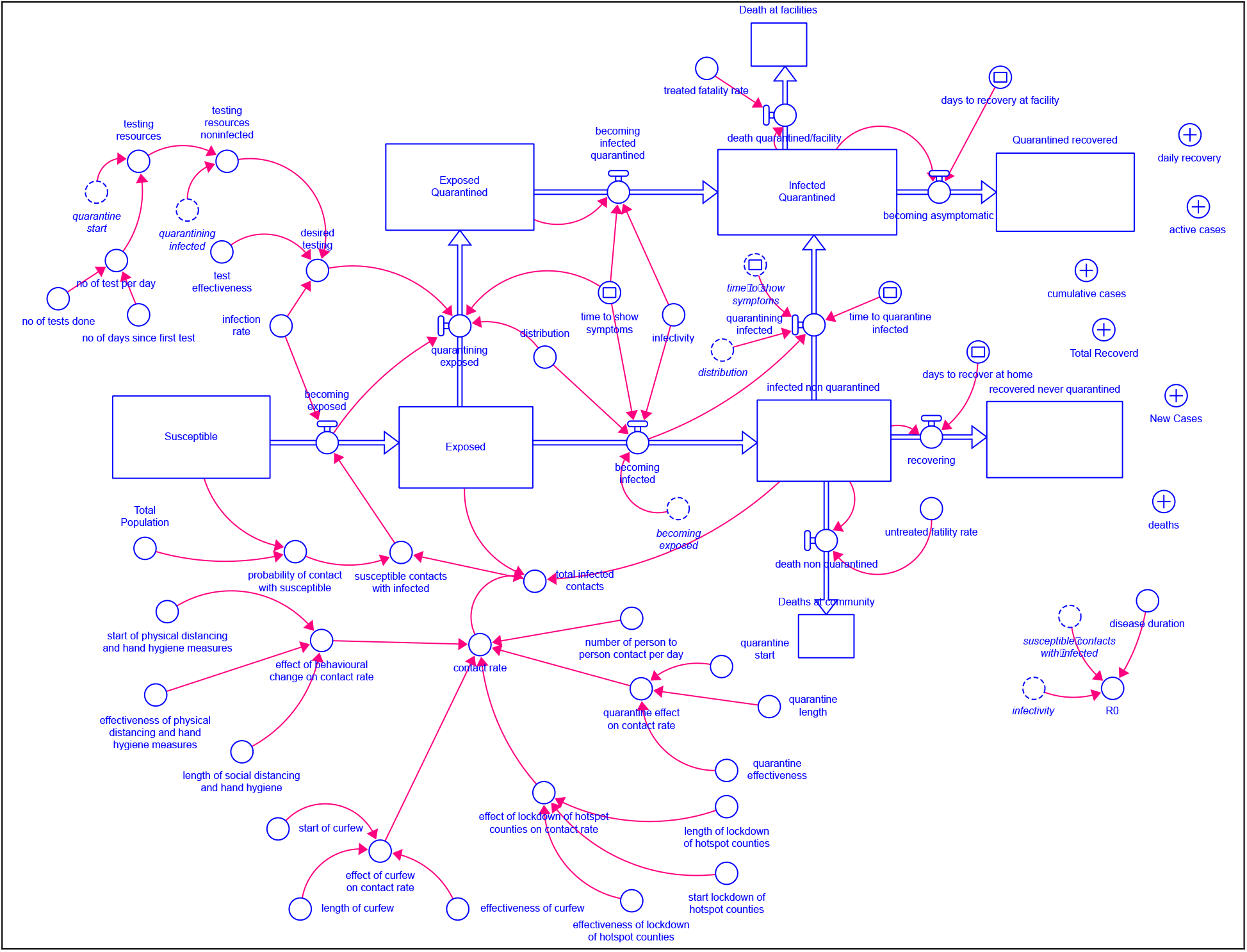
Overview of Stock and Flow diagram of Covid-19 trajectory in Kenya.

Figure 2 shows the overview of the stock and flow diagram of possible Covid-19 pathways in Kenya. From the stocks and flow diagrams, two possible pathways are explored. The exposed, infected and never tested individuals progress through the various stages of Covid-19 disease states and may never visit a health facility. While those who get tested for the disease transit through the same pathways but managed under an institution, health facility or structured home-based care.

Figure 3 is a screenshot of the interphase window showing nobs and numeric input slides that can be used to vary inputs into the model. Figure 3 also shows the screenshot of current scenario of current covid-19 management approaches in Kenya.

**Fig 3:**
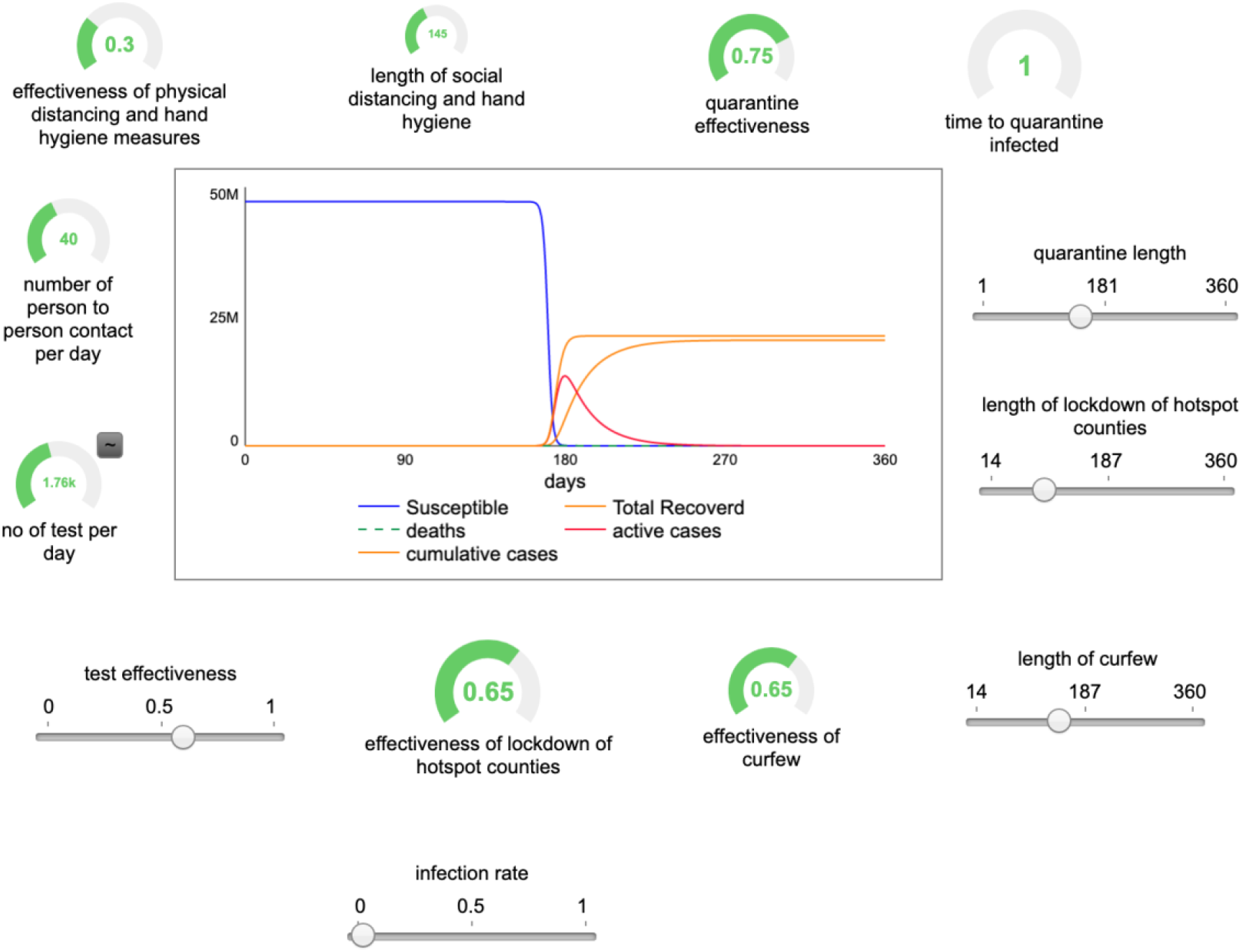
Screen shot of Interphase page showing the possible trajectory of current state (base-case scenario) of Covid-19 Interventions in Kenya.

### 2.2 Model Calibration

We used published data to calibrate the model and where data was not available, expert opinion was sought. We simulated the model for Population of 47.2 million people representing the Kenya Population.

The basic parameters used to seed are presented in Table 1 and these represent the base case scenario. The information used to verify the model structure was sourced form SEIRD publications[28][30], recent case studies on Covid-19, World Health Organizations(WHO), Our World in Data, Kenyan Government press reports and expert judgement.

**TABLE 1:** LIST OF VARIABLES USED AS BASE CASE SCENARIO AND TO SEED THE MODEL.

| Variable | Value | Reference | Comment |
| --- | --- | --- | --- |
| **GENERAL POPULATION SECTOR** |  |  |  |
| Date of First COVID 19 case reported | 13<sup>th</sup> March 2020 | MoH guidelines(Ministry of Health, 2020a) |  |
| Susceptible population/Kenya Population | 47.5 Million | Brand et.al, (2020), Forecasting the scale of the COVID-19 epidemic in Kenya<br>https://www.medrxiv.org/content/10.1101/2020.04.09.20059865v1.full.pdf |  |
| Incubation time | 5 days on average | (Lauer et al., 2020)(Singhal, 2020) |  |
| Disease duration | Range from 1-14 days | (Singhal, 2020)(Lauer et al., 2020) |  |
| Fraction requiring hospitalization | 20%<br>(80% of the cases are mild) | https://www.health.go.ke/wp-content/uploads/2020/06/Updated-Case-Management-Guidelines-26_03_20-1.pdf |  |
| R<sub>0</sub> | 2.4-3.3 | (Park, Cook, Lim, Sun, & Dickens, 2020) |  |
| Base Contact Rate/ persons per day | 40 | We approximated 15 in the rural areas and 65 in urban areas. |  |
| Case Fatality Rate | 0.5-3.5%<br>(2.5% facility)<br>(2.9% for community) | (Njenga et al., 2020)(Harries, Martinez, & Chakaya, 2020)<br>https://www.health.go.ke/wp-content/uploads/2020/06/Kenya-SITREP-089-14-Jun-2020.pdf |  |
| Number of cases reported at start | 1 | 13<sup>th</sup> March 2020 |  |
| Date of start of Quarantine | 10 | Press Brief. Directive given on the 10<sup>th</sup> day since confirmation of first case in Kenya. |  |
| Average number of days of quarantine | 16 | MoH guidelines, inclusive of two days for discharge logistics |  |
| Length of self/ facility Quarantine policy | Ongoing Since first day of government directive |  | Simulated for 145 days as per time of this model development |
| Effectiveness of Quarantine | 65% | Expert opinion, | Data not available |
| Rate of Infection | 1.9 % | (Yancy, 2020)(Bendavid et al., 2020)(Nopsopon et al., 2020) | We adopted the Rate of China |
| Rate of Deaths at Home | 2.5/1000 | Expert opinion, estimated to be lower that of Hospital facility Rate | Data not available |
| Time taken to recover at home | 16 days | Estimated. (If recovery with treatment may reduce recovery time with 2 days, then recovery at home may increase with 2 days | Data not available |
| Time taken to recover at Health Facilities | Average 10 days<br>(7-12 days) | (Beigel et al., 2020)(Wang et al., 2020)(Hung et al., 2020) | Based in clinical improvement after some pharmaceutical's treatment |
| Number of available tests per day/ base testing resources | Average 1760per day | Calculate as the total number of tests done/number of days since first test | The ministry has reported challenges in acquiring test kits thus inconsistencies in testing |
| Effectiveness of tests | 60% | (Hung et al., 2020) |  |
| Start of lockdown of 4 hotspots counties | 25 days | Press release | 6<sup>th</sup> April 2020 |
| Length of lockdown of 4 hotspot counties | 92 days | Lockdown Terminated after 92 days through a Press Release |  |
| Effectiveness of lockdown of 4 hotspot counties | 65% | Expert opinion |  |
| Curfew start | 15 days since reporting of first case in Kenya | Press release |  |
| Curfew effectiveness | 75% | Expert Opinion |  |
| Curfew length | 145 days | Continuous since first announced |  |
| Social distancing and hand washing start | 11 days | Expert Opinion, the population took almost two weeks before serious start of hand washing |  |
| Effectiveness of social distancing and hand washing | 30% | Expert opinion,<br>Our world in data less than 30% of Kenyan population had access to hand washing facilities. |  |
| Length of social distancing and hand washing | 120 days | Expert Opinion, the population took almost two weeks before serious start of hand washing |  |
| Normal contact rate/ number of people to person touch per day | 40 per day | Expert Opinion<br>(Approx. 15 per day in rural setting and 65 in urban setting) |  |
| Quarantine effectiveness | 70% | (Hung et al., 2020) 48.7%<br>Expert Opinion higher than Hung as it was forced<br>quarantining as a government directive. | Based on ABS of influenza |
| Quarantine length | 16 days | Average reported date by various patients on informal<br>bases |  |

The base case scenario represents the current status of Covid-19 interventions in Kenya. We performed model adjustments by varying auxiliary variables associated with various interventions in Covid-19 management. The first day on each output was taken as the 13^th^ March 2020, when Kenya confirmed its first case of Covid-19.

### 2.3 Assumptions of the model

The model assumes:

1. A homogenous and static population hence the effect of new births and immigration was excluded. The stocks are therefore conserved.
2. That movement across compartments/stocks is a given time step
3. Incubation period of 5 days
4. That new cases can be detected on testing
5. Untested cases will not be identified; however, they progress through stocks in a similar way as the detected cases while in the community.
6. That the unidentified cases in the community may get ‘opportunistic testing’ and follow the pathways of detecting cases in corresponding stocks.
7. That there is conferred immunity after recovery.
8. Lockdown, curfew, social distancing and hygiene measures, curfew and quarantining will have an impact on the number of people to person contacts per day.
9. That physical distancing includes closure of schools, churches and other social gatherings including adaptation of public transport to Covid-19 guidelines by the government.
10. That it would be possible for populace to consciously taken note and reduce the number of person to person contact per day.
11. Since the infection from the virus is reinforcing, and therefore have an exponential growth, the measures taking to limit its transmission are expected to have a counterbalancing effect on its growth.

### 2.4 Model Validation and Simulation

Model validation done through a walk and passed adequacy and extreme conditioning tests. The ability to replicate historical Covid-19 data in Kenya was also demonstrated.

We simulated the model for an initial period of 365 days and used 0.0625 DT and RK4 integration method. The differential equations for the complete model are available in equation mode of the Stella model (Supplementary file 1). We built up different scenarios through varying the variables. We varied the effectiveness of selected variables (physical distancing and hand hygiene, curfew, and quarantining) according to the recommended WHO guidelines of 50%, 80% and 95% [31]. We added a ‘realistic’ effectiveness of the selected variables defined as what the experts felt were achievable levels of interventions in the country. The realistic coverage was operationalized as physical distancing and handwashing hygiene at 65%, effectiveness of curfew at 80%, quarantining at 80% and person to person contact at 30 per day.

The effectiveness of lockdown of hot spot counties was not varied as lifting of the lockdown of selected counties happened in the course of developing this model. However, the impact of the lockdown was included in the base-case scenario.

We analyzed the impact of varying levels of effectiveness against trends on possible active infections, deaths and the number of days saved from pushing the peak of Covid-19 infection. The ability of the model to reproduce historical data was assessed through comparative runs from raw data reported on our world in data and simulated run at base case scenario

## 3.0 Results

The ability to reproduce historical data is shown in Figure 4 demonstrating the comparison between the two curves of daily reported cases and simulation results from our model. The two curves are similar in shape even though the Simulated numbers are higher due to low testing levels of Kenya, hence possibility of missed cases.

**Fig 4.**
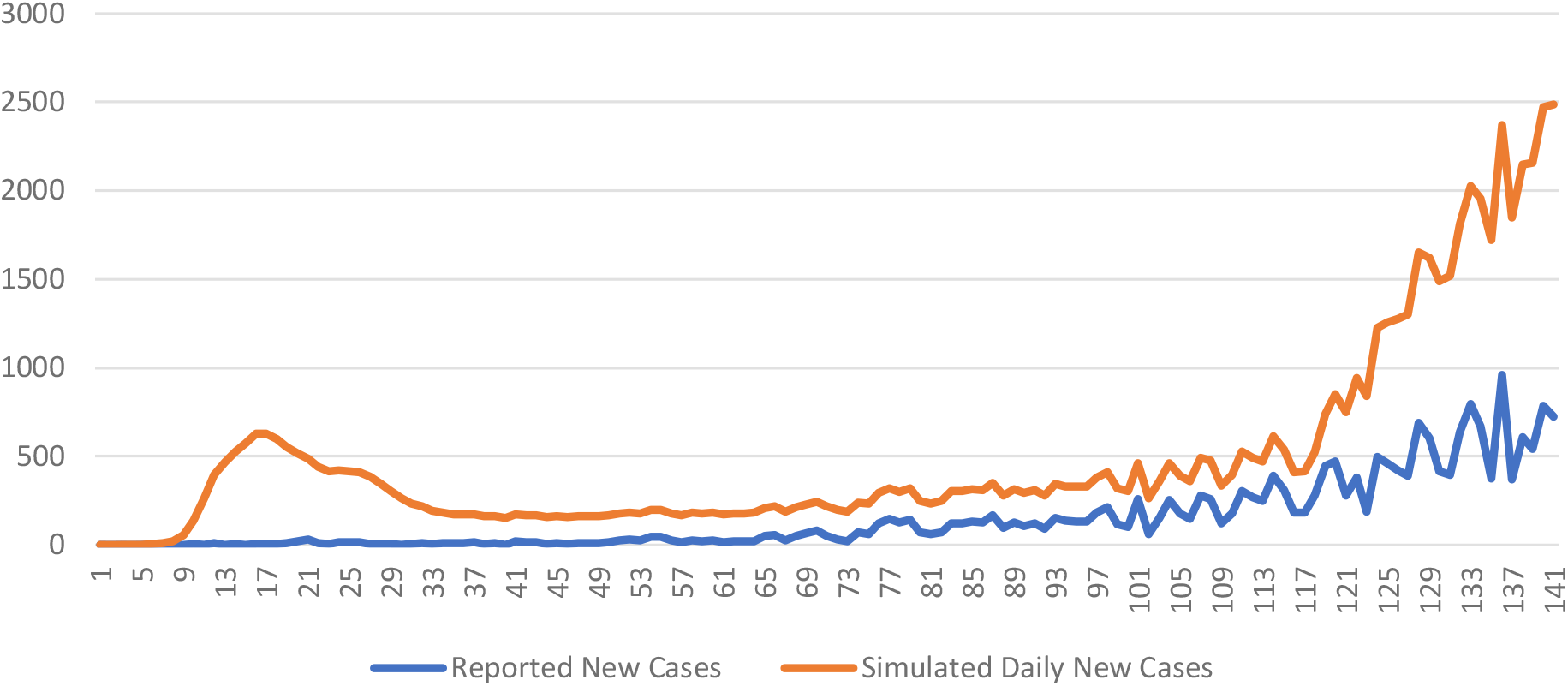
Comparison of Reported Daily New Cases and Simulated Daily New Cases

Kenya has a rate of positive cases at 13% by end of July 2020 (Our World in Data, 2020) meaning that country performed many tests relative to the size of the outbreak, thus many cases are likely to be unreported.

Similarly, Fig 5 compares the results of cumulative Cases of raw data and base case scenario run. The curve is similar but the numbers are higher due to possible low levels of testing.

**Fig 5:**
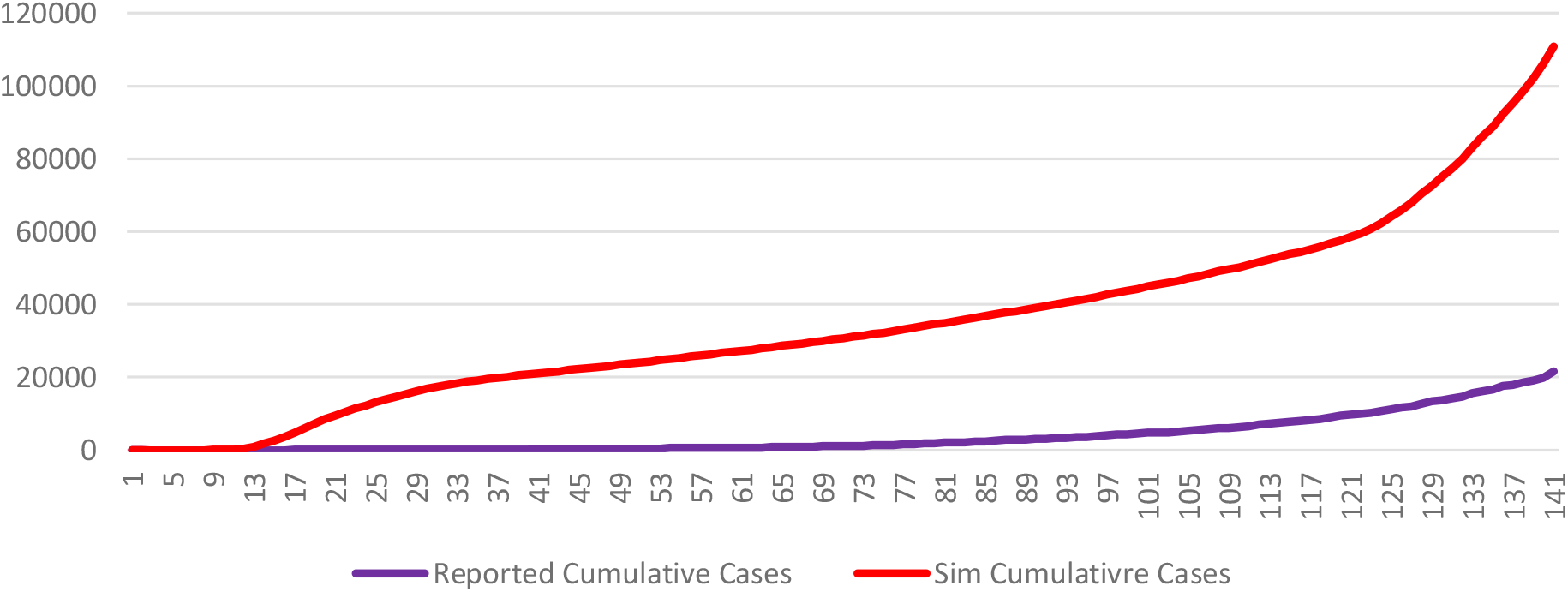
Comparison of Cumulative Reported Cases and Simulated Cumulative Cases for the First 145 Days

The base case scenario (with current interventions) is presented in Figure 6. Showing that the peak infections of Covid-19 are likely to occur on day 178 (September 2020) with approximately 13.6 million active cases and 34,000deaths. We compared the Base case scenario through varying effectiveness of selected non pharmaceutical interventions.

**Figure 6.**
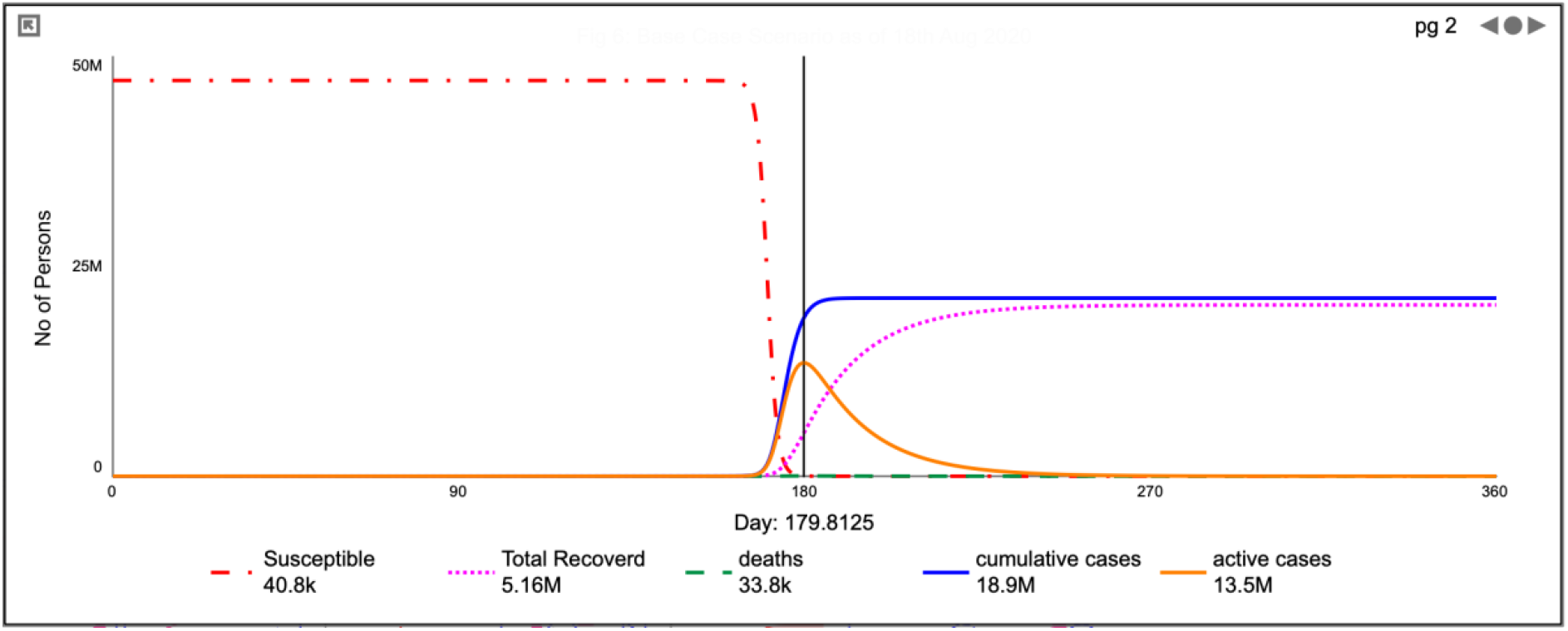
Base Case scenario of Covid-19 possible trajectory in Kenya as of 18^th^ August 2020.

### 3.1 Physical Distancing and Hand Hygiene

All activities geared towards physical distancing as well as hand hygiene measures included all behavioral adaptation such as closure of social gatherings and modification of transport systems among others. Holding all other variables as at base case scenario, we varied the levels of effectiveness of physical distancing at the WHO recomemded levels and the effectiveness levels the experts felt were realistic or could be achievable. The impact of various levels of behavioural adaptation were simulated and compared to base case scenario. The results are demonstrated in Figure 7 a and 7b shows that 50%, 65%(realistic level), 80% and 95% levels of effectiveness would push the peak of Covid-19 with 2, 4 and 6 days respectively with a minimal reduction in both active cases and deaths.

**Fig 7a.**
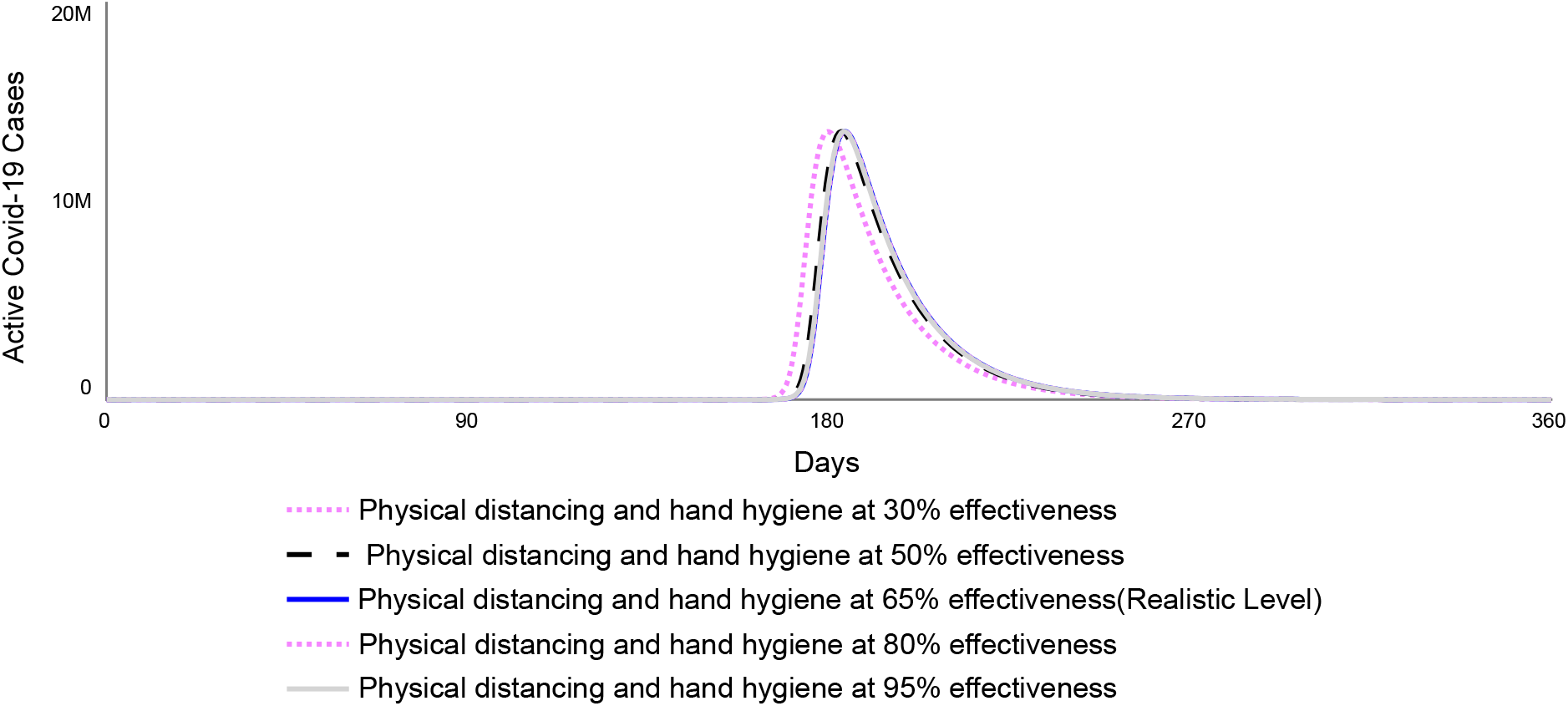
Impact of Physical Distancing and Hand Hygiene on Active cases

**Fig 7b.**
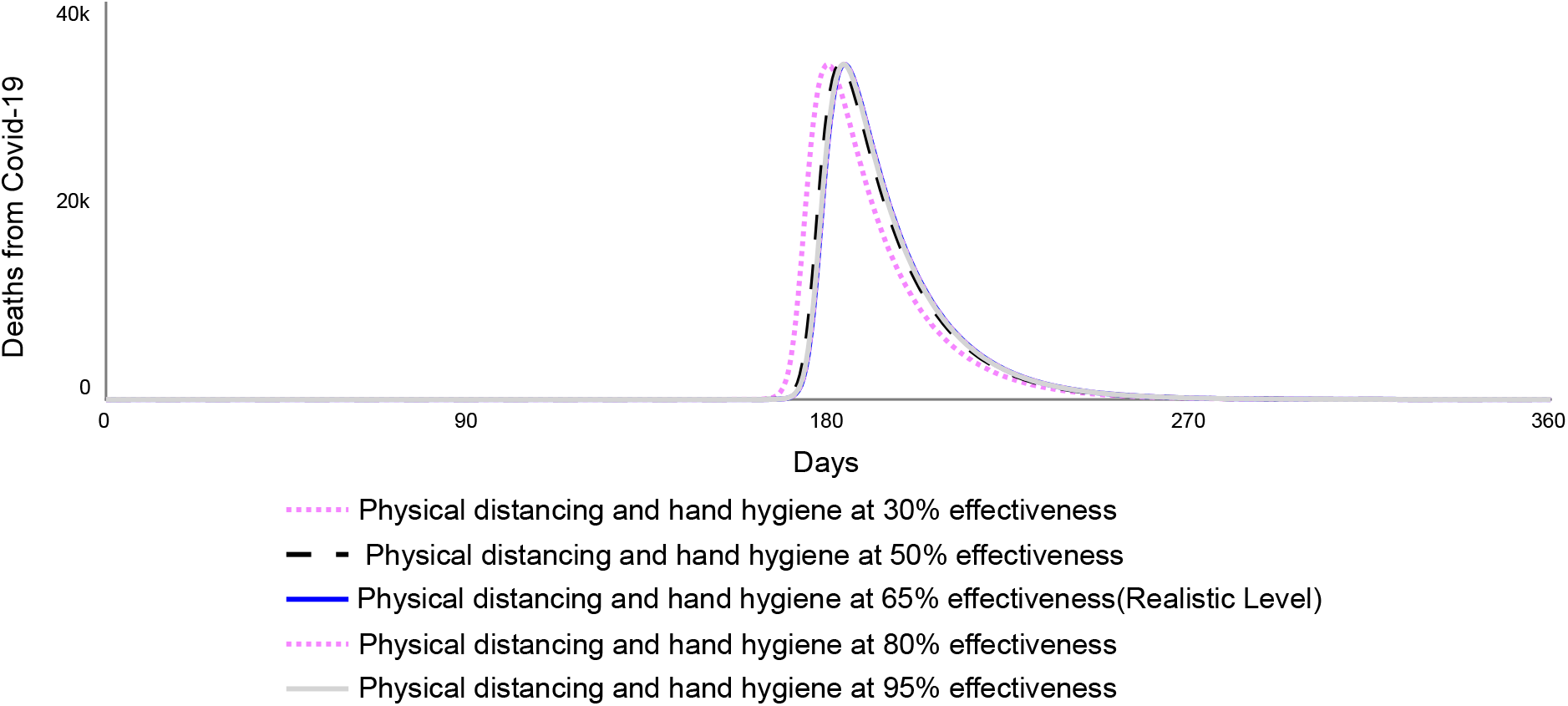
Impact of Physical distancing and hand Hygiene on Mortality

### 3.2 Impact of Movement Restrictions; Curfew and Lockdown of Hot Spot Counties

Movement restriction in Kenya was implemented in two ways. A national wide curfew and lockdown of 5 hotspot counties. The lockdown was modified as it involved closure of borders of the hotspot counties as well as some sub-sections of two(Nairobi and Mombasa) of the hotspot counties. A curfew was effected for the first time on the 15^th^ day since confirmation of first Covid-19 case in Kenya. In the first month, the curfew was effected from 7pm to 5am daily. This was later varied to 9pm to 4am daily. By the time of publication, the 9pm to 4am curfew was ongoing. We did not differentiate this varying of curfew timings in our model. The experts felt that the 7pm to 5am curfew was more effective at approximately 75% while generally people tended to take the 9pm to 4 am curfew less seriously with a suggested effectiveness of 65%. Overall, the base-case effectiveness of curfew was estimated at 65%. We varied the effectiveness of curfew at the 80% and 95% WHO levels and adopted the 80% effectiveness as our realistic level.

Since the lockdown of hotspot counties had been lifted by the time of development of this model, we did not vary the variables associated with lock down. The effect of lockdown was however included in the base case scenario.

As demonstrated in Fig 8 a and Fig 8 b, curfew may have resulted to shifting the peak of both active cases and deaths from 178 days at base case to 183 days at 95% effectiveness with minimal reduction in the number of active cases as well as the number of deaths.

**Fig 8a.**
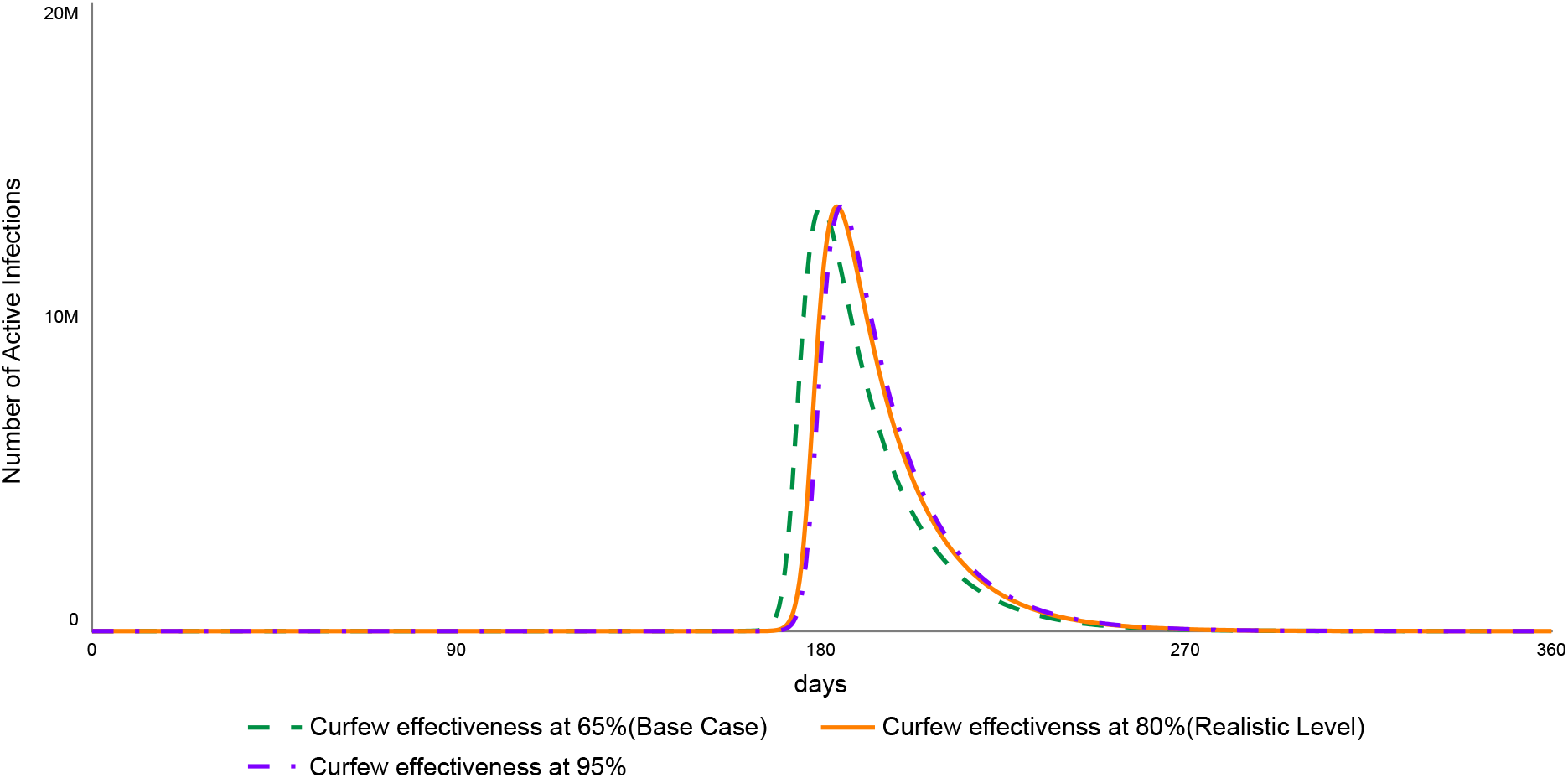
Impact of curfew effectiveness on active cases

**Fig 8b.**
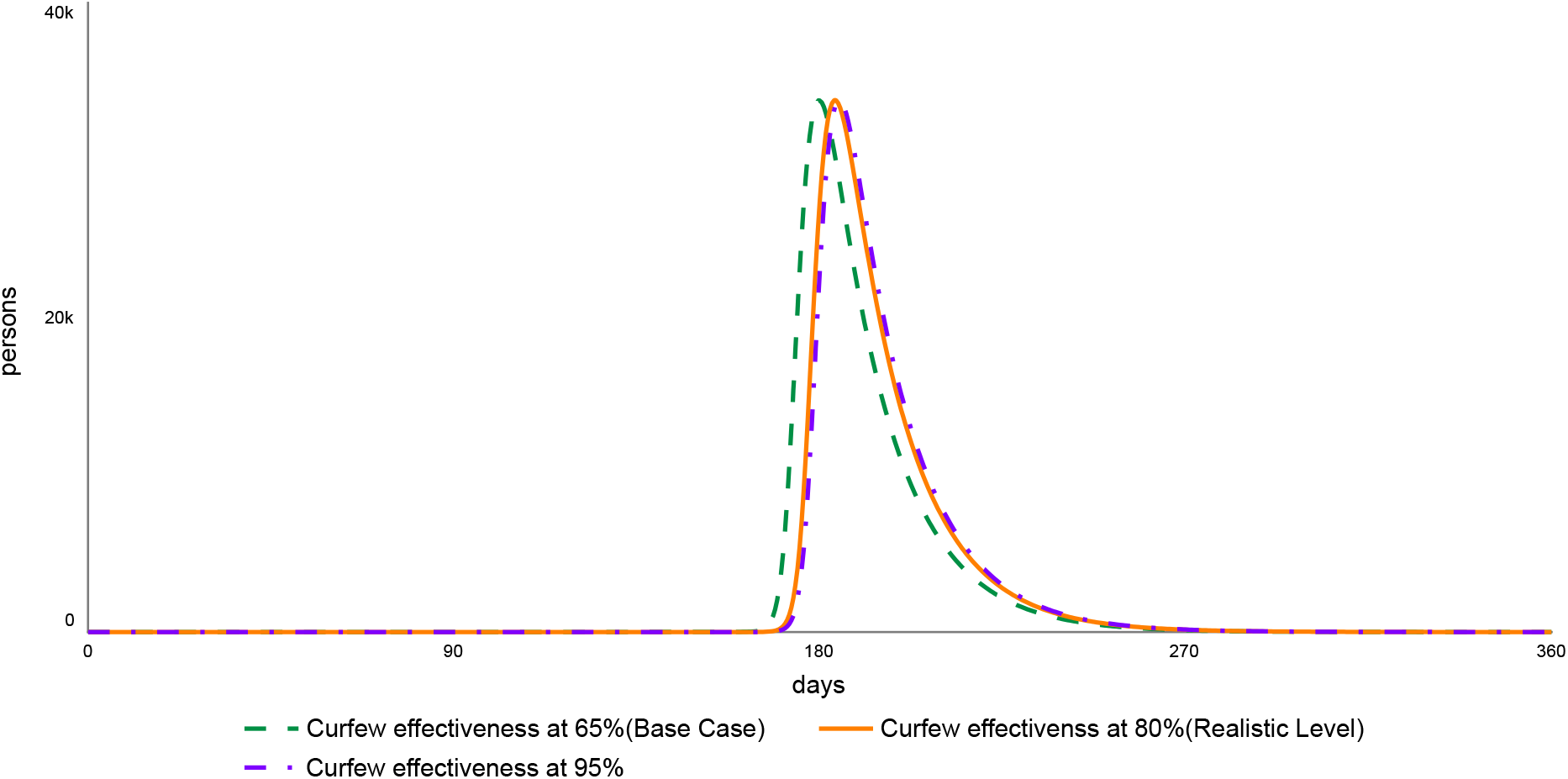
Impact of varying levels of curfew effectiveness on deaths from Covid 19

### 3.3 Impact of Quarantine on Covid-19 in Kenya

The initial approach to quarantining was self-quarantining of any persons who came in to the country after the Covid-19 pandemic was declared. This was found not to be effective and by the of day 21, the government implored compulsory quarantine for new entrants into the country. Special government designated facilities were utilized. Each quarantined person was expected to cater for their own cost of quarantining with the cheapest facility charging $20 per day. This led to an uproar from the general population, and there were reported cases of a few people escaping from some quarantine facilities. The quarantine facilities were viewed and reported by the local print and media as an avenue of perpetuating police brutality. Quarantine in Kenya was therefore a challenge. After about 4 weeks from the reporting of the first Covid-19 case in Kenya, the government resulted to free quarantine at government own centers. We estimated the overall effectiveness of quarantine at 75%. We held all other variables as at base case scenario and varied the effectiveness of quarantining. As demonstrated in Fig 9a and 9b increasing the effectiveness of quarantining from 75% to 80% and 95% would push the peak of active cases and deaths with 3.5 days and 5.5 days respectively.

**Fig 9a.**
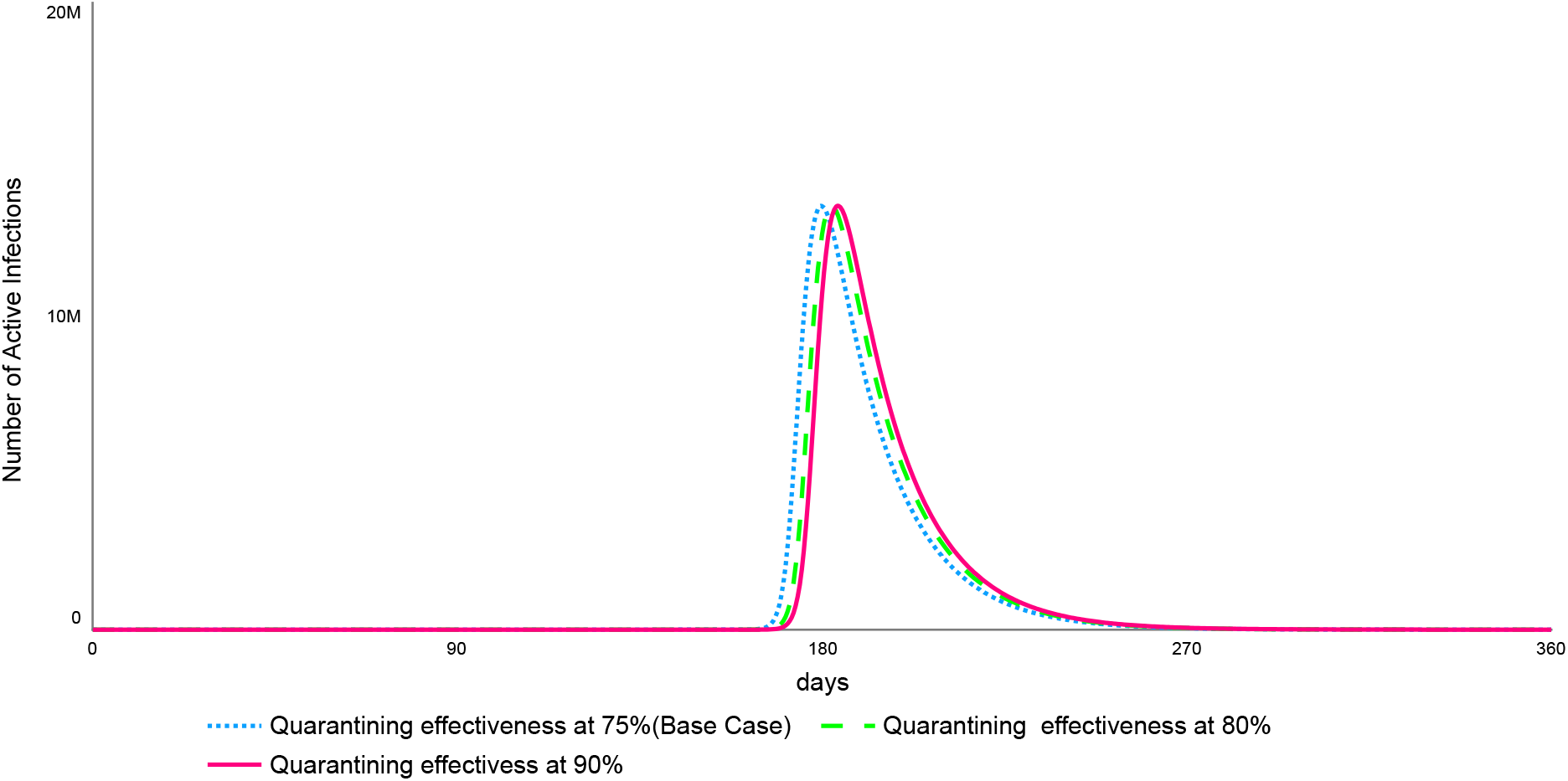
Impact of quarantining effectiveness on active cases

**Fig 9b.**
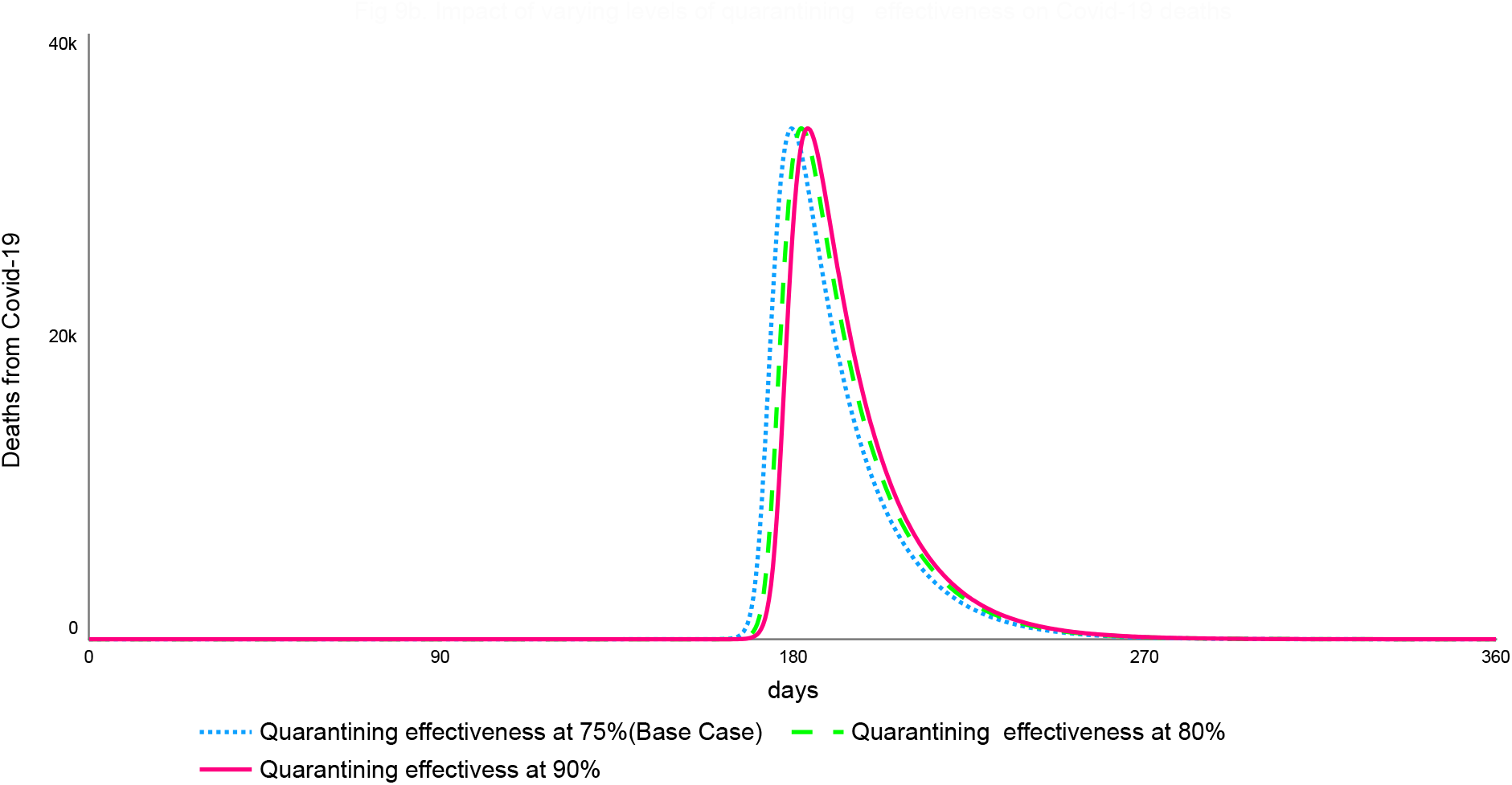
Impact of varying levels of quarantining effectiveness on Covid-19 deaths

### 3.4 Number of person to person contact per day

Since total lockdown of the country would not be feasible due to possible catastrophic social-economic impact, we assumed a scenario whereby the general population would make a conscious effort to interact with a maximum specific number of people at most per day. This would have various impact on Covid-19 trajectory. We estimated the number of person to person contact per day to be at 65 in urban centers and 15 in rural areas thus an average of 40 person to person contacts per day. Holding all other variables as at base case scenario, we varied this person to person contact per day by 50, 40 and 30 resulting to the possible peak of covid-19 cases at 173^rd^, 179 ^th^ and 226^th^ days respectively. We adapted 30 person to person contact per day as our realistic coverage. A reduction from 40 to 30 person to person contact per day would move the peak of Covid-19 cases with approximately 21 days.

### 3.5 Realistic Scenario

The realistic scenario was what we felt the Country had potential to achieve in Covid-19 management. As demonstrated in Fig 11, realistic intervention levels of all the selected non pharmaceutical interventions; effectiveness of physical distancing and hand hygiene at 65%, curfew at 80%, quarantining at 80% and person to person contact at maximum of 30) would result to a delay of peak of Covid-19 cases from 178^th^ day since first confirmed infection to a peak of 246^th^ day allowing approximately 67 extra days for preparedness of health care system. The new peak would likely be late November to mid-December 2020.

**Fig 10:**
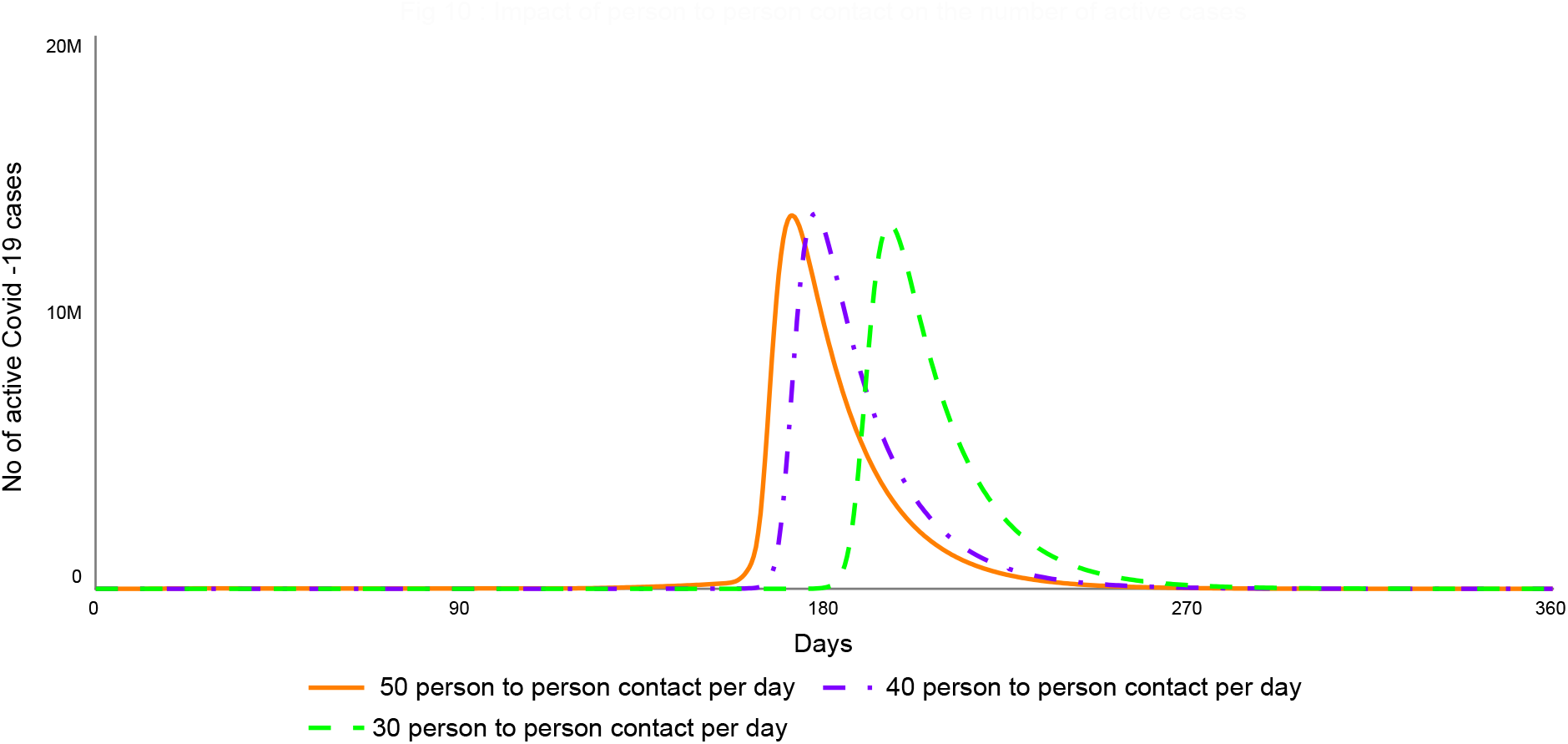
Impact of person to person contact on the number of active cases

**Fig 11.**
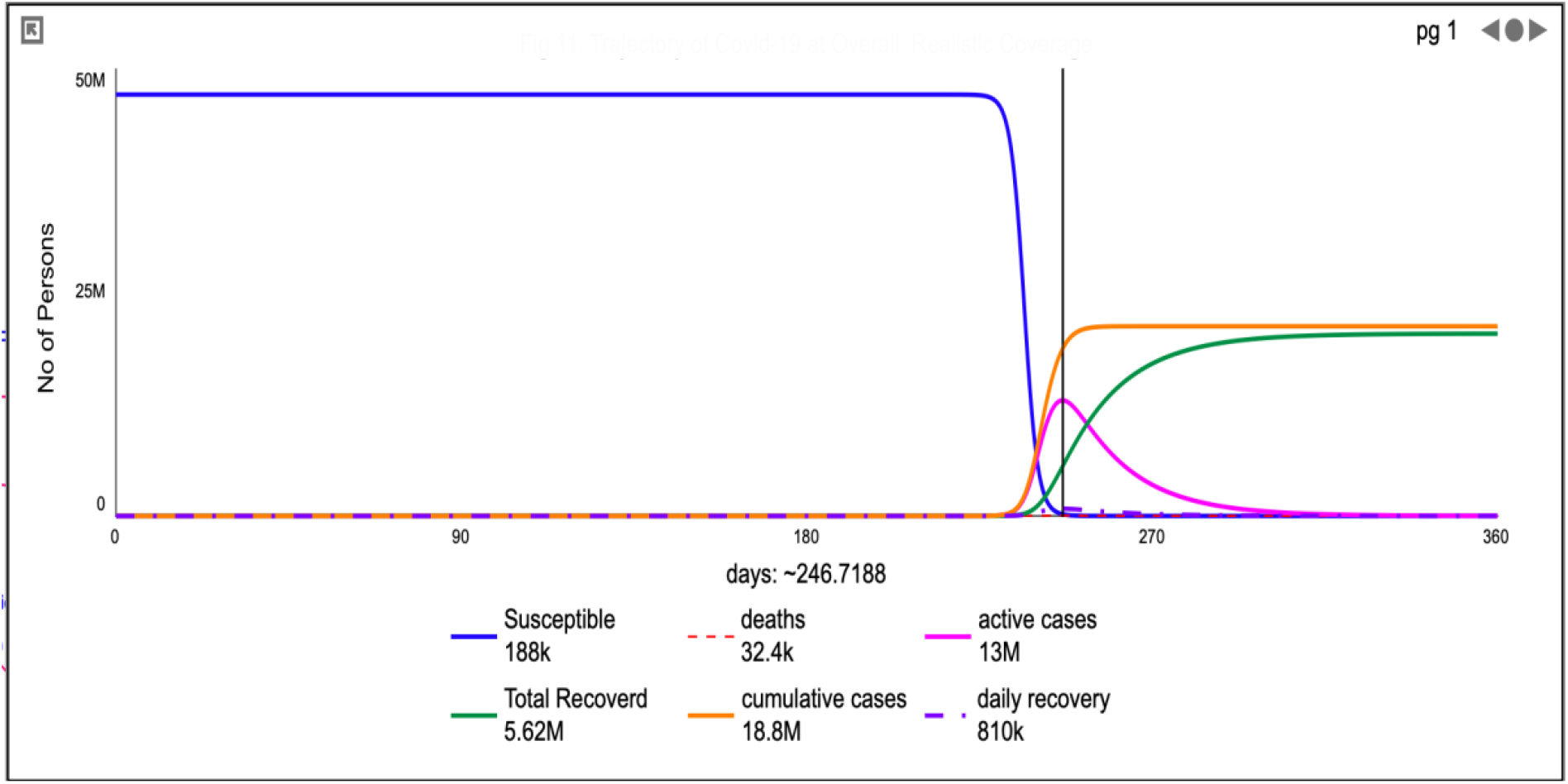
Trajectory of Covid-19 at Overall Realistic Coverage

## 4.0 Discussion

In this paper, we simulated the effect of non-pharmaceutical measures on the progression of Covid-19 pandemic in Kenya. Our model used locally adapted non-pharmaceutical measures for managing the spread of Covid-19 and flattening the curve. We ran the model using the WHO reported coverage rates of 50%, 80% and 95%, for modelling interventions in a pandemic (We assumed that the general population would make a conscious effort to interact with a specific number of people at most per day) We simulated several scenarios starting with current status quo of the pandemic interventions in Kenya and varied the scenarios based on the WHO coverage proportions and realistic coverage in Kenya. The model provided results of three scenarios: the base scenario that represents the current population coverage of interventions in Kenya (hand hygiene and physical distancing coverage of 30%, curfew 65%, border closures of 30% and quarantine 75%); the second scenario where we used the coverage rates in the base scenario but varied the rates for hand hygiene and physical distancing with WHO expected coverage of 50%, 80%, and 95% consecutively; and the third scenario, our realistic model, that represents physical distancing and hand hygiene coverage of 65%; 30 contact persons per day, curfew coverage of 80% and quarantine of 80%. The performance of each of the models was measured in terms of the number of days the proposed scenario would delay the peaks of the infection and mortality rates; overall reduction in the number of infections, and the number of days saved to the flat the pandemic curve in Kenya.

Our simulated base scenario was effective in replicating the current data on the ground. When we compared this model to the actual data on the number of new cases reported daily, there was a similar trend (figure 1). Our simulation did however show a high number of new cases are being missed. On average, for every 3 cases that were reported there are seven other cases that go undetected (ratio 3:10) (figure 4). Similar findings have been reported elsewhere with possible reasons being insufficient testing kits, difficulties with contact tracing due to limited resources (Kobia and Gitaka 2020) and stigma resulting in failure to present for testing (Turner-Musa et al. 2020) Further, our model predictions of the current status quo (base scenario) show that over 50% of the Kenyan population will be infected with the virus by six months of the pandemic (figure 1).

Our simulation suggest that multiple and feasible interventions need to be adopted to limit the spread of the virus and flatted the curve. When we simulated single interventions such as physical distancing and hand hygiene, even with a population uptake of this intervention at 95%, the infection and mortality peak rates could only be extended with 3.5 and 5.5 days respectively.

Our suggested scenario depicting realistic intervention levels of all the selected non pharmaceutical interventions would delay of peak by 67 days allowing a modest amount of time to prepare the mitigate possible overwhelming of the health care system. The new peak would then be late November to mid-December 2020. While controlling for physical distancing and hand hygiene, the greatest impact was on the extending the curve was seen when person to person contact was varied

### 4.1 Implications for the application of the model

The ideal scenario would be for Kenya to achieve a population uptake of 100% for all the suggested measures to control the pandemic. However, the social acceptability and feasibility of such level coverage in a resource limited setting like Kenya that has populous cities, overcrowded housing, high usage of public transport is dismal[33].

Majority of the hotspot countries are overcrowded and access to soap and water, and hand sanitizers remains a challenge too with alternative hygiene measures being fronted [34]. We therefore predict that use of realistic model, would allow the government time to organize resources to deal with the mortality and infection peaks.

### 4.2 Limitations of the study

Covid-19 is dynamic and the data may vary drastically. Our model is based on person to person contact and provides suggestions that take into account the current situation in the country. The application of the model may be limited to Kenya because the mixing patterns of individuals may differ in other regions and countries and across cultures. While we acknowledge sufficient data was used to populate the model, we also leave room for incorporating new knowledge to further refine the model. We also did not classify the severity of Covid-19 cases.This model does not attempt to predict the course of Covid-19 in Kenya but rather generates hypothesis as to possible Covid-19 Trajectories from possible non-pharmaceutical interventions.

## 5.0 Conclusion

The current non-Pharmaceutical interventions are likely to have pushed the peak date of Covid-19 cases to September 2020.Enhanced intervention would push this peak by Approx. 67 days giving extra time for the preparedness health systems. A realistic combination of non -pharmaceutical interventions may have greater impact on Covid-19 Trajectories in Kenya.

A simplified language of the number of person to person contact per day may be a more understandable message. SDM is a useful tool in seeking a deeper understanding of impact of non-pharmaceutical interventions in Covid-19.

## Data Availability

All data used is available is detailed. The Stella Model can be availed on request from the corresponding author.

## Acknowledgments

We acknowledge the contribution of Robert Eberlein through the Webinar of Modeling the COVID-19 Pandemic: A Primer and Overview, in Isee Systems. We appreciate the invaluable support of Dr Jonathan Moizer and Prof Jonathan Lean of University of Plymouth and Danny Ibarra-Vega of IRCACS(Columbia).

## Funding Acknowledgements

This research received no specific grant from any funding agency in the public, commercial, or not-for-profit sectors.

